# The excess insulin requirement in severe COVID-19 compared to non-COVID-19 viral pneumonitis is related to the severity of respiratory failure and pre-existing diabetes

**DOI:** 10.1101/2020.10.03.20206284

**Authors:** Sam M. Lockhart, Harry Griffiths, Bogdan Petrisor, Ammara Usman, Julia Calvo-Latorre, Laura Heales, Vishakha Bansiya, Razeen Mahroof, Andrew Conway Morris

## Abstract

**Objective:** Severe COVID-19 has been anecdotally associated with high insulin requirements. It has been proposed that this may be driven by a direct diabetogenic effect of the virus that is unique to SARS-CoV-2, but evidence to support this is limited. To explore this, we compared insulin requirements in patients with severe COVID-19 and non-COVID-19 viral pneumonitis.

**Research Design:** Retrospective cohort study of patients with severe COVID-19 admitted to our intensive care unit between March and June 2020. A historical control cohort of non-COVID-19 viral pneumonitis patients was identified from routinely collected audit data.

**Results:** Insulin requirements were similar in patients with COVID-19 and non-COVID-19 viral pneumonitis after adjustment for pre-existing diabetes and severity of respiratory failure.

**Conclusions:** In this single center study, we could not find evidence of a unique diabetogenic effect of COVID-19. We suggest that high insulin requirements in this disease relate to its propensity to cause severe respiratory failure in patients with pre-existing metabolic disease.

## Introduction

There is emerging interest in the effect of COVID-19 on glucose homeostasis, with some suggesting that COVID-19 may be a uniquely diabetogenic virus [1]. Observations advanced to support this hypothesis include anecdotal reports of high insulin requirements in patients with severe COVID-19 [2-5]. However, to date there has been no longitudinal study of patients with COVID-19 to empirically assess insulin requirements. We sought to address this deficiency by comparing insulin requirements in patients admitted to intensive care with severe COVID-19 to a historical control group with severe non-COVID-19 viral pneumonitis.

## Research Design and Methods

### Case Identification

Patients admitted to ICU in Addenbrooke’s Hospital, Cambridge with a positive SARS-CoV-2 test were identified. Patients were deemed to have COVID-19 pneumonitis and included in the study if they were SARS-CoV-2 positive and were treated for respiratory failure that was attributed to COVID-19 pneumonitis by the treating team during their ICU stay.

The non-COVID-19 viral pneumonitis cohort was identified from the ICU’s routinely collected audit data from 2014 to 2019. Patients admitted with a diagnosis of viral pneumonia recorded as their primary, secondary or ultimate reason for admission to intensive care were included in the analysis.

### Data Collection

Data collection was undertaken manually from electronic medical records by trained, clinical investigators. Blood glucose data was collected using proprietary analytics software, Qlikview. Further detail is available in the supplementary appendix.

### Matching

Patients admitted with COVID-19 pneumonitis were matched according to diabetes status and level of respiratory support received in a 1:1 ratio to a historical control cohort of patients admitted to ICU with non-COVID-19 viral pneumonitis. Further data is available in the supplementary appendix.

### Statistical Analysis

Details of the statistical analysis are available in the supplementary appendix.

### Ethical review statement

As a retrospective analysis of anonymized, routinely collected data the requirement for ethical committee review and consent was waived by the National Health Service Health Research Authority, the evaluation was registered with the responsible healthcare organization (Cambridge University Hospitals NHS Foundation Trust).

## Results

During the study period March to June 2020 we identified 92 patients treated for severe COVID-19 within our intensive care unit (Supplementary Appendix Figure 1). In univariate linear regression analysis, BMI, pre-existing Type 1 or Type 2 Diabetes and severity of respiratory failure assessed by ordinal scale (0 – self ventilating, 1-mechanical ventilation, 2 – neuromuscular blockade, 3 – nebulized epoprostenol, 4 – prone ventilation, 5 – extracorporeal membrane oxygenation) were all associated with insulin dose when analyzed by the average dose across the whole ICU stay or by the highest cumulative insulin dose over a 24 hour period (Supplementary Appendix, Table 1).

To determine if the observed insulin requirements were a unique feature of COVID-19 we examined insulin requirements in 46 patients admitted to our ICU with a diagnosis of non-COVID-19 viral pneumonitis between 2014 and 2019. 89% of the non-COVID-19 cohort had a confirmed viral isolate consistent with viral pneumonia. 5 patients included in the analysis did not have any relevant positive virology related to their ICU admission but were diagnosed with viral pneumonitis on clinical grounds while 2 patients had more than 1 virus identified that could have explained their symptoms (Supplementary Appendix, Table 2 for a breakdown of viral isolates). Insulin requirements were higher in patients with COVID-19 versus non-COVID-19 viral pneumonitis while mean glycemic indices were comparable in both groups (Figure 1A-B and Supplementary Appendix Figure 2A-B). However, the groups were poorly balanced with respect to diabetes status and requirement for respiratory salvage therapies (Supplementary Appendix, Table 3), multiple regression analysis did not identify any association between COVID-19 and insulin dose after adjustment for pre-existing diabetes and severity of respiratory failure (Supplementary Appendix Table 4). Similarly, when COVID-19 and non-COVID-19 viral pneumonitis patients were matched (1:1, N = 36 per group) by the level of respiratory support required and pre-existing diabetes status, the proportion of patients requiring insulin (COVID-19: 41%, non-COVID-19: 49%) and insulin dose was similar in those who were treated with insulin (Figure 1C-D and Supplementary Figure 2C-D).

**Figure 1.**
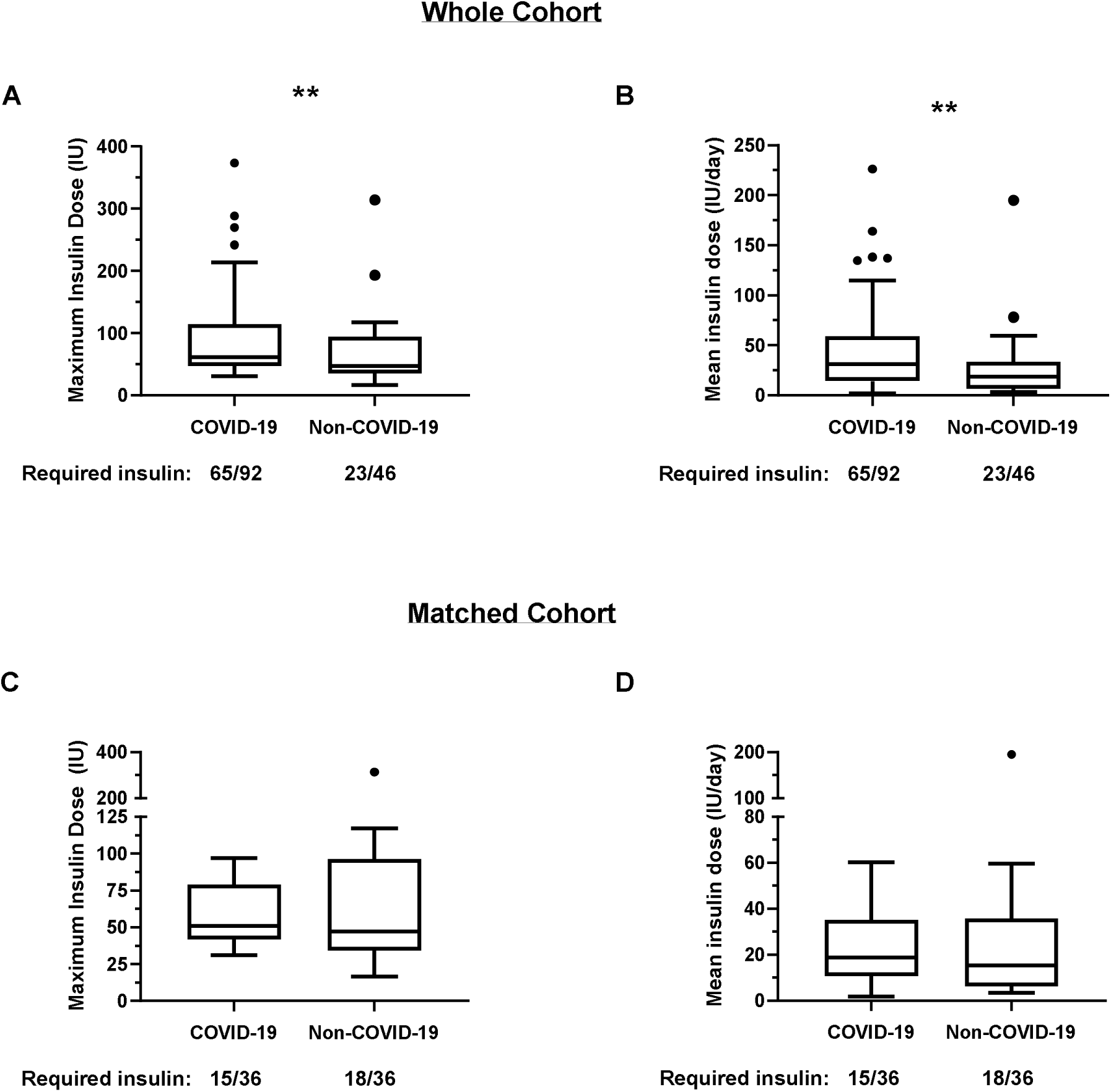
Insulin requirements of patients admitted to ICU with COVID-19 and non-COVID-19 viral pneumonitis. **A)** Tukey box plots of maximum cumulative insulin requirements in a single day in the complete cohort, P = 0.003 by Mann-Whitney U test. **B)** Tukey box plots of mean daily insulin dose day in the complete cohort, P = 0.003 by Mann-Whitney U test **C)** Tukey box plots of maximum cumulative insulin requirements in a single day in the matched cohort, P = 0.46 by Wilcoxon signed rank test. D) Tukey box plots of mean daily insulin dose day in the matched cohort, P = 0.52 by Wilcoxon signed rank test. For clarity only insulin dose for patients receiving insulin plotted (i.e. no zero values plotted) but statistical tests are performed on the whole dataset.

Finally, we sought to determine if the insulin requirements observed in patients with COVID-19 resolved in convalescence. We had information on discharge medication in 61/62 patients who survived to hospital discharge. Of these patients 39 required insulin while in ICU but on discharge only 5 patients were discharged from hospital on subcutaneous insulin, 4 of whom were on insulin prior to admission. The one patient commenced on insulin had Type 2 Diabetes prior to admission managed with oral therapy, insulin was discontinued soon after discharge when they were readmitted with hypoglycemia. There was no initiation or escalation in oral hypoglycemic agents in the cohort, however we cannot comment on the adequacy of glycemic control. Similar findings from a smaller cohort have recently been reported [3].

## Conclusions

We report here, to the best of our knowledge, the first longitudinal assessment of glycemia and insulin requirements in patients who are critically ill with COVID-19. Our data suggest that high insulin requirements observed in patients with severe COVID-19 is related to the severity of respiratory failure and pre-existing diabetes mellitus rather than a direct diabetogenic effect of SARS-CoV-2. Therefore, there is currently no rationale to adopt novel strategies or different glycemic thresholds to manage hyperglycemia in COVID-19 compared to other viral pneumonias. Moreover, the comparable insulin requirements in COVID-19 and non-COVID-19 viral pneumonitis when severity of illness and pre-existing diabetes are accounted for argue against a direct action of SARS-CoV-2 on the pancreatic β-cell as the likely cause of hyperglycaemia in COVID-19, as has been suggested previously [6]. In addition, our data suggest that the hyperglycemia observed in patients with COVID-19 is transient and resolves in convalescence and leads us to caution studies describing new-onset diabetes in patients with COVID-19 based on cross-sectional blood glucose measurements [7].

However, our analysis is limited by its relatively small sample size and its single center nature. Further determination of the specific effects of COVID-19 infection as opposed to the effects of general critical illness will require collection of data from larger, well matched cohorts with other precipitating causes of stress hyperglycemia. It should be noted that we cannot exclude the existence of rare hyperglycemic syndromes triggered by SARS-CoV-2 infection. For example, our study cannot inform the etiological role of SARS-CoV-2 infection in autoimmune diabetes as has been studied in larger epidemiological analyses [8-10]. Large, registry-based studies (e.g. COVIDIAB registry) with prolonged follow-up will be required to study this and to determine the long-term effects of COVID-19 infection on glucose metabolism more broadly.

In conclusion, high insulin requirements in severe COVID-19 likely relate to the severity of respiratory failure and the high prevalence of metabolic disease in patients with severe illness. When these factors are accounted for, insulin requirements are comparable to that seen in non-COVID-19 viral pneumonitis.

## Data Availability

Fully anonymised data is available on request to the corresponding author.

## Author Contributions

SL, VB, RM and ACM conceived and designed the study. SL, HG, BP, AU, LH collected the data. SL and ACM wrote the manuscript and performed the analysis. All authors reviewed the manuscript for important intellectual content. ACM is the guarantor of this work.

## Funding

SL is supported by a National Institute of Health Research Academic Clinical Fellowship. ACM is supported by a Clinical Research Career Development Fellowship from the Wellcome Trust (WT 2055214/Z/16/Z)

## Disclosures

The authors have no relevant conflict of interests to declare.

## Supplementary Appendix

### Methods

#### Study Centre

All patients included in the study were managed in Addenbrooke’s hospital intensive care unit which includes a 20-bed general intensive care unit with a further 12 high and intermediate dependency beds which were repurposed as critical care beds as required during the pandemic, and a 27-bed specialist neurocritical care unit.

#### Glycemic control

Our departmental policy for management of hyperglycaemia in critical care advises commencement of a variable rate intravenous insulin therapy when two consecutive blood glucose readings are >10 mmol/L if there has been no recent (within 6 hours) hypoglycaemia. The rate of intravenous insulin infusion is adjusted by bedside nursing staff depending on blood glucose according to a standard prescription which can be titrated by medical staff with the aim of maintain a target blood glucose of 6-10mmol/L. In patients with persistent hyperglycaemia or high insulin requirements, subcutaneous basal insulin may be commenced under the advice of the specialist diabetes team. When this was added it was solely with the intention of improving glycaemic control. There was not, to our knowledge, any case where subcutaneous insulin therapy was used in an effort to spare syringe pumps during the COVID-19 pandemic.

#### Data Collection

Demographic data was manually collected from review of the participant electronic medical record. Admission values for cross sectional variables such as age and BMI were recorded.

A diagnosis of pre-morbid diabetes mellitus was recorded if the patient was on any anti-hyperglycemic therapy, if they had a recorded diagnosis of diabetes mellitus in their problem list or if they had a pre-morbid HbA_1c_ that was diagnostic of Diabetes according to World Health Organisation Diagnostic Criteria (48 mmol/mol or over). Diabetes mellitus was attributed to Type 2 Diabetes unless there was a positive diagnosis of Type 1 Diabetes recorded in the notes. Prior to admission hyperglycaemic therapy was documented according to the reconciled medications list in the electronic medical record and the medicines administration record was used to determine anti-hyperglycaemic therapies used while in hospital. The discharge summary was used to determine antihyperglycaemic medications that were continued on discharge.

Insulin doses recorded were the sum of all subcutaneous and intravenous insulin administered during the patient’s stay in intensive care.

Acute corticosteroid use was examined and the number of days that corticosteroids were administered was recorded. We did not include corticosteroid use that was prescribed regularly prior to admission.

APACHE II scores were collected on admission to intensive care and were obtained from routinely collected audit data.

#### Matching

Patients admitted with COVID-19 pneumonitis were matched in a 1:1 ratio to a historical control cohort of patients admitted to ICU with non-COVID-19 viral pneumonitis. Subjects were strictly matched so that the level of respiratory support received (e.g. mechanical ventilation, neuromuscular blockade, epoprostenol therapy, prone ventilation and extra-corporeal membrane oxygenation) and diabetes status were identical. If there was more than one suitable match then age was used (as age was significantly associated with insulin requirements when added to the model presented in supplementary table 3), if age was similar (tolerance +/- 5 years) then BMI was reviewed, if BMI was similar (tolerance +/-2.5 kg/m^2^) then Sex was reviewed. These criteria were pre-specified prior to matching and undertaken blinded to knowledge of insulin requirements.

#### Statistical analysis

Student’s t-test was used for comparing means of normally distributed data, Mann-Whitney U test and Wilcoxon Rank test were used for comparing unpaired and paired non-normally distributed data, respectively.

The level of maximum respiratory support required during the ICU stay was assigned an ordinal scale : 0 – self ventilating, 1-mechanical ventilation, 2 – neuromuscular blockade, 3 – nebulized epoprostenol, 4 – prone ventilation, 5 – extracorporeal membrane oxygenation. The regression co-efficients and p-values reported are from a model encoding respiratory support as a continuous variable but a sensitivity analysis encoding respiratory support as a categorical variable showed similar results.

For the purposes of analysis, the number of days receiving steroids was normalized according to the length of stay to avoid the confounding effects of duration of ICU stay.

Insulin requirements (both maximum use in a single day and mean daily use) were right skewed and were squareroot transformed to facilitate regression analysis. For multiple regression analysis diabetes status and level of required respiratory support were included in the model as these two variables were unbalanced in the COVID-19 and non-COVID-19 viral pneumonitis groups and were the most significant predictors of insulin requirement. The addition of age and BMI to the model (which were both significant in univariate analysis) did not improve the predictive ability of the model as assessed by a likelihood ratio test, nor did they alter the relationship between COVID-19 and insulin requirement.

The results reported are from linear regression analysis of the whole data set but similar results were observed (e.g. COVID-19 was not a significant predictor of insulin requirements after addition of an index of respiratory support and diabetes status were added to the model) using tobit regression with censoring at 0 or if insulin requirement was dichotomized to those requiring and not requiring insulin and analyzed by binomial regression.

Statistical analysis was conducted using Graphpad Prism version 8.4.3 and R.

**Supplementary Figure 1.**
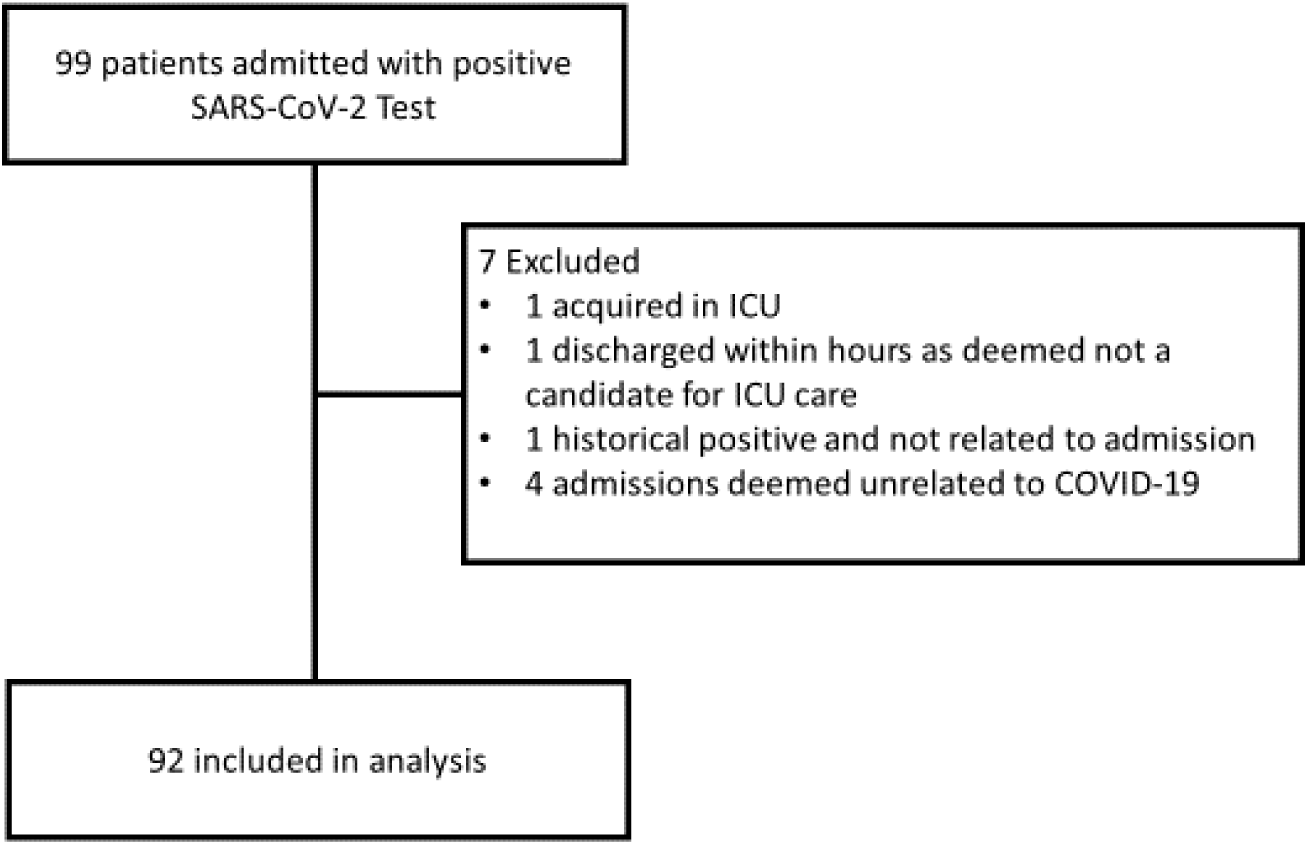
Consort diagram illustrating patient inclusion

**Supplementary Figure 2.**
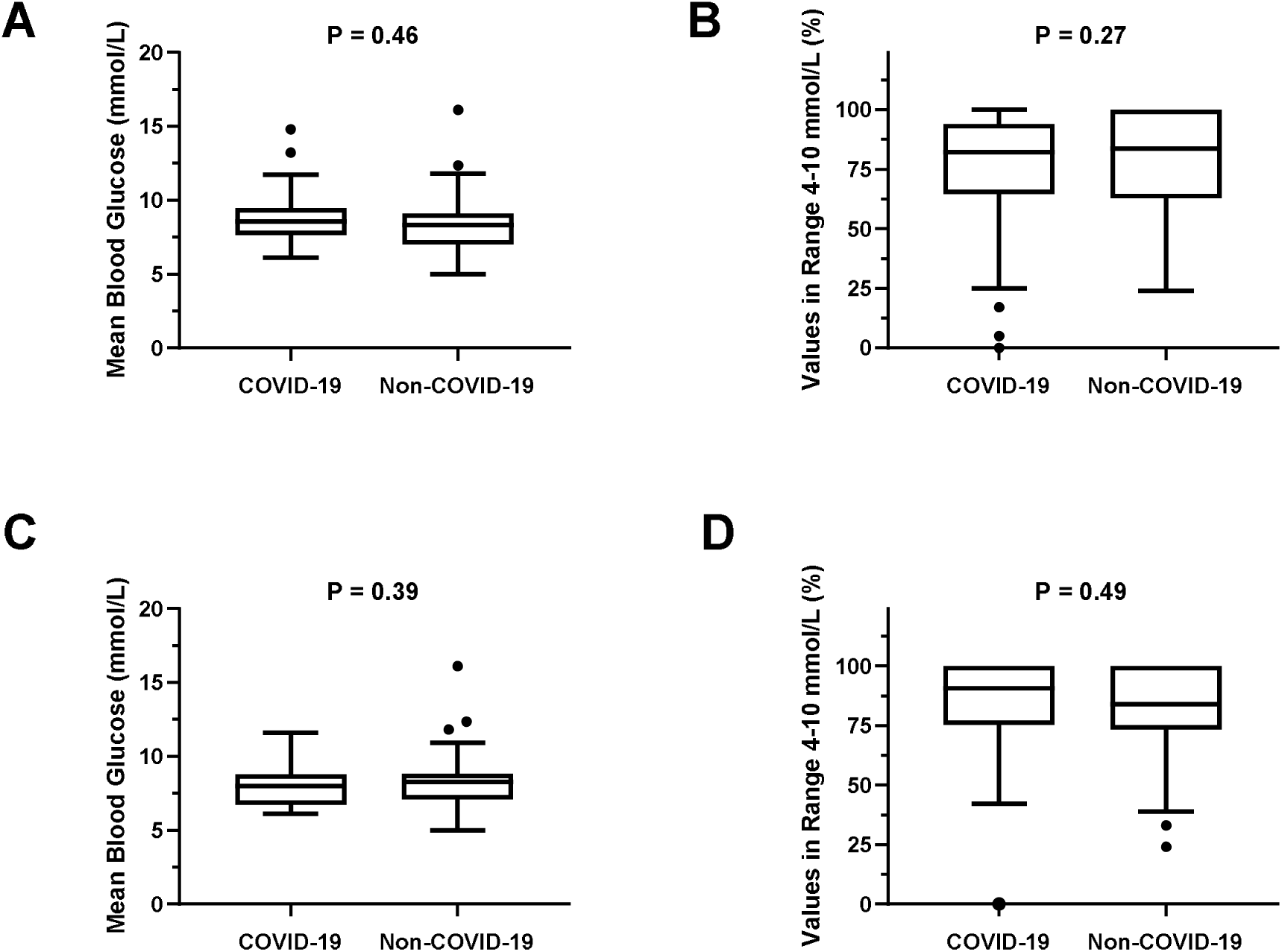
Glucose levels of patients of patients admitted to ICU with COVID-19 and non-COVID-19 viral pneumonitis. **A)** Tukey Box plots of mean blood glucose in the complete cohort, P = 0.42 by t-test **B)** Tukey Box plots of percentage time in target range (4-10mmol/L) in the complete cohort, P = 0.27 by Mann-Whitney U test **C)** Tukey Box plots of mean blood glucose in the matched cohort, P = 0.39 by paired t-test. **D)** Tukey Box plots of percentage time in target range (4-10mmol/L) in the matched cohort, P = 0.49 by Wilcoxon signed rank test

**Supplementary Table 1.** Correlation coefficients +/- standard error, multiple R-squared and p-values from univariate linear regression analysis of maximum insulin requirement in a single day and mean insulin requirements per day, following a square root transformation.

|  | **Beta** | **R<sup>2</sup>** | **P-Value** |
| --- | --- | --- | --- |
| **Maximum insulin requirement in a single day** |  |  |  |
| **Age** | 0.08 +/- 0.04 | 0.05 | 0.03 |
| **Male** | -0.23 +/-1.10 | 0.0004 | 0.83 |
| **Type 1 Diabetes** | 7.58 +/- 3.42 | 0.04 | 0.03 |
| **Type 2 Diabetes** | 5.60 +/- 1.05 | 0.23 | <0.001 |
| **BMI** | 0.19 +/- 0.08 | 0.06 | 0.02 |
| **APACHE II** | 0.19 +/- 0.09 | 0.04 | 0.05 |
| **Respiratory Support** | 1.64 +/- 0.26 | 0.30 | <0.001 |
| **Steroid Exposure** | 1.37 +/- 1.80 | 0.006 | 0.76 |
| **Mean insulin requirements per day** |  |  |  |
| **Age** | 0.04 +/- 0.03 | 0.02 | 0.14 |
| **Male (n)** | -0.32 +/- 0.82 | 0.02 | 0.70 |
| **Type 1 Diabetes** | 6.34 +/- 2.52 | 0.07 | 0.01 |
| **Type 2 Diabetes** | 4.53 +/- 0.75 | 0.29 | <0.001 |
| **BMI** | 0.16 +/- 0.06 | 0.08 | 0.008 |
| **APACHE II** | 0.12 +/- 0.07 | 0.03 | 0.08 |
| **Respiratory Support** | 1.10 +/- 0.20 | 0.25 | <0.001 |
| **Steroid Exposure** | 1.33 +/- 1.33 | 0.01 | 0.32 |

**Supplementary Table 2.** Viral isolates in the Non-COVID-19 viral pneumonitis cohort.

| **Viral Isolate** | **N** |
| --- | --- |
| Flu A | 22 |
| Flu B | 4 |
| Enterovirus | 2 |
| VZV | 1 |
| CMV | 1 |
| HMPV | 2 |
| Rhinovirus | 3 |
| Parainfluenza | 6 |
| Non-Typeable picornavirus | 1 |
| Adenovirus | 1 |
| No viral isolate – clinical diagnosis | 5 |

**Supplementary Table 3.**
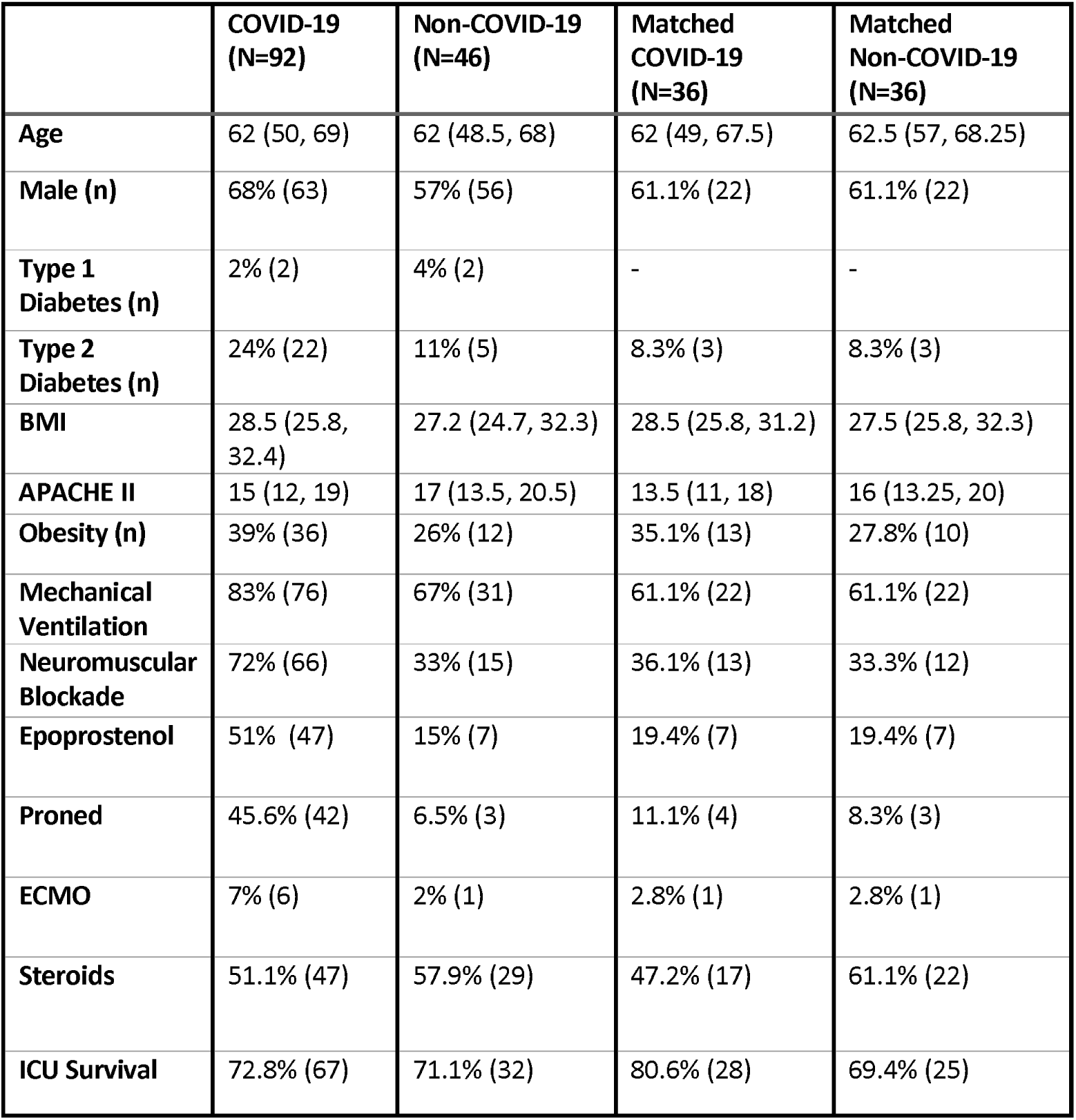
Demographics of patients with COVID-19 viral pneumonitis and non-COVID-19 viral pneumonitis in the whole cohort and in the cohort matched for diabetes status and severity of respiratory failure. Categorical variables are summarized as % (n) and continuous variables as median (Q1, Q3).

**Supplementary Table 4.** Correlation coefficients +/- standard error, multiple R-squared and P-values from multivariable linear regression analysis of maximum insulin requirement in a single day and mean insulin requirements per day, following a square root transformation.

|  | **Beta** | **P-Value** |
| --- | --- | --- |
| **Maximum insulin requirement in a single day, $R^2 = 0.47$** |  |  |
| **COVID-19** | 0.10 +/- 0.71 | 0.89 |
| **Diabetes Mellitus** | 4.66 +/- 0.75 | <0.001 |
| **Respiratory Support** | 1.41 +/- 0.21 | <0.001 |
| **Mean insulin requirements per day, $R^2 = 0.50$** |  |  |
| **COVID-19** | 0.20 +/- 0.50 | 0.71 |
| **Diabetes Mellitus** | 5.60 +/- 1.05 | <0.001 |
| **Respiratory Support** | 1.64 +/- 0.26 | <0.001 |

## Notes

### Competing Interest Statement

The authors have declared no competing interest.

### Clinical Trial

Not registered as study was a retrospective analysis of routinely collected data

